# Genetic Liability to Cannabis Use Disorder and COVID-19 Hospitalization

**DOI:** 10.1101/2020.11.15.20229971

**Authors:** Alexander S. Hatoum, Claire L. Morrison, Evan A. Winiger, Emma C. Johnson, Arpana Agrawal, Ryan Bogdan

**Affiliations:** Washington University School of Medicine, Department of Psychiatry; Institute for Behavioral Genetics, University of Colorado Boulder; Washington University in St. Louis, Department of Psychological & Brain Sciences

**Keywords:** Cannabis Use Disorder, Genome-wide association statistics, COVID-19, genetic correlations, quasi-experimental design, tobacco use

## Abstract

Behavioral and life style factors plausibly play a role in likelihood of being hospitalized for COVID-19. Genetic vulnerability to hospitalization after SARS-CoV2 infection may partially relate to comorbid behavioral risk factors, especially the use of combustible psychoactive substances. Paralleling the COVID-19 crisis has been increasingly permissive laws for recreational cannabis use. Cannabis Use Disorder (CUD) is a psychiatric disorder that is heritable and genetically correlated with respiratory disease, independent of tobacco smoking. By leveraging genome-wide association summary statistics of CUD and COVID-19, we find that at least 1/3^rd^ of the genetic vulnerability to COVID-19 overlaps with genomic liability to CUD (rg=.34, p=0.0003). Genetic causality as a potential mechanism of risk could not be excluded. The association between CUD and COVID-19 remained when accounting for genetics of trying marijuana, tobacco smoking (ever smoking regularly, cigarettes per day, smoking cessation, age of smoking initiation), BMI, fasting glucose, forced expiration volume, education attainment, and Townsend deprivation index. Heavy problematic cannabis use may increase chances of hospitalization due to COVID-19 respiratory complications. Curbing excessive cannabis use may be an essential strategy in COVID-19 mitigation.

## Introduction

Paralleling the Coronavirus Disease 2019 (COVID-19) pandemic has been an increase in substance use (1) and a continuation of increasingly permissive laws surrounding cannabis. On November 3^rd^, 2020, American voters in 4 U.S. states (Arizona, Montana, New Jersey, South Dakota) voted to join 11 that have already legalized recreational cannabis use. Legalization is associated with increased use and 20% of individuals who have tried cannabis develop cannabis use disorder (CUD) (2), a moderately heritable (50-60%) psychiatric syndrome that shares genetic risk with respiratory disease (3). As the heterogeneous presentation of COVID-19 is partially attributable to host genomic background and respiratory symptoms are the primary reason for hospitalization and death (4), genomic liability to CUD may contribute to severe COVID-19 presentation.

## Results

We leveraged genome-wide association study (GWAS) summary statistics to estimate whether genetic liability to CUD (n case=14,080, n control=343,726) (3) may plausibly influence COVID-19 hospitalization (n case=6,492, n population control=1,012,809) (4). *First*, Linkage Disequilibrium Score Regression (5) analyses revealed that genetic liability for CUD and COVID-19 hospitalization are correlated (SNP-r_g_=0.34, p=0.0003, and remained significant after Bonferroni correction for pairwise genetic correlations across all traits; **Figure 1**). *Second*, genetic liability to tobacco smoking phenotypes was also shared with COVID-19 hospitalization (**Figure 1**). A series of genomic structural equation models (gSEM) (6) revealed an independent genomic association between CUD and COVID-19 hospitalization (**Table 1**) when accounting for genomic liability to tobacco phenotypes (i.e., cigarettes per day, age of becoming a regular smoker, ever being a regular smoker, smoking cessation(7)), lifetime cannabis use(8), cardiometabolic traits (i.e., BMI (9), fasting glucose (10)), respiration (i.e., 1-second forced expiratory volume (11)) and socioeconomic status (i.e., educational attainment (12), Townsend deprivation index (11)). The only tobacco phenotype that retained a significant independent association in these models was smoking cessation, which was protective (**Table 1**). *Third*, and finally, latent causal variable (13) (LCV) analyses provided evidence that liability to CUD may be genetically causal for COVID-19 hospitalization (genetic causality proportion estimate = 0.63(.21), p=4.0e-6).

**Figure 1.**
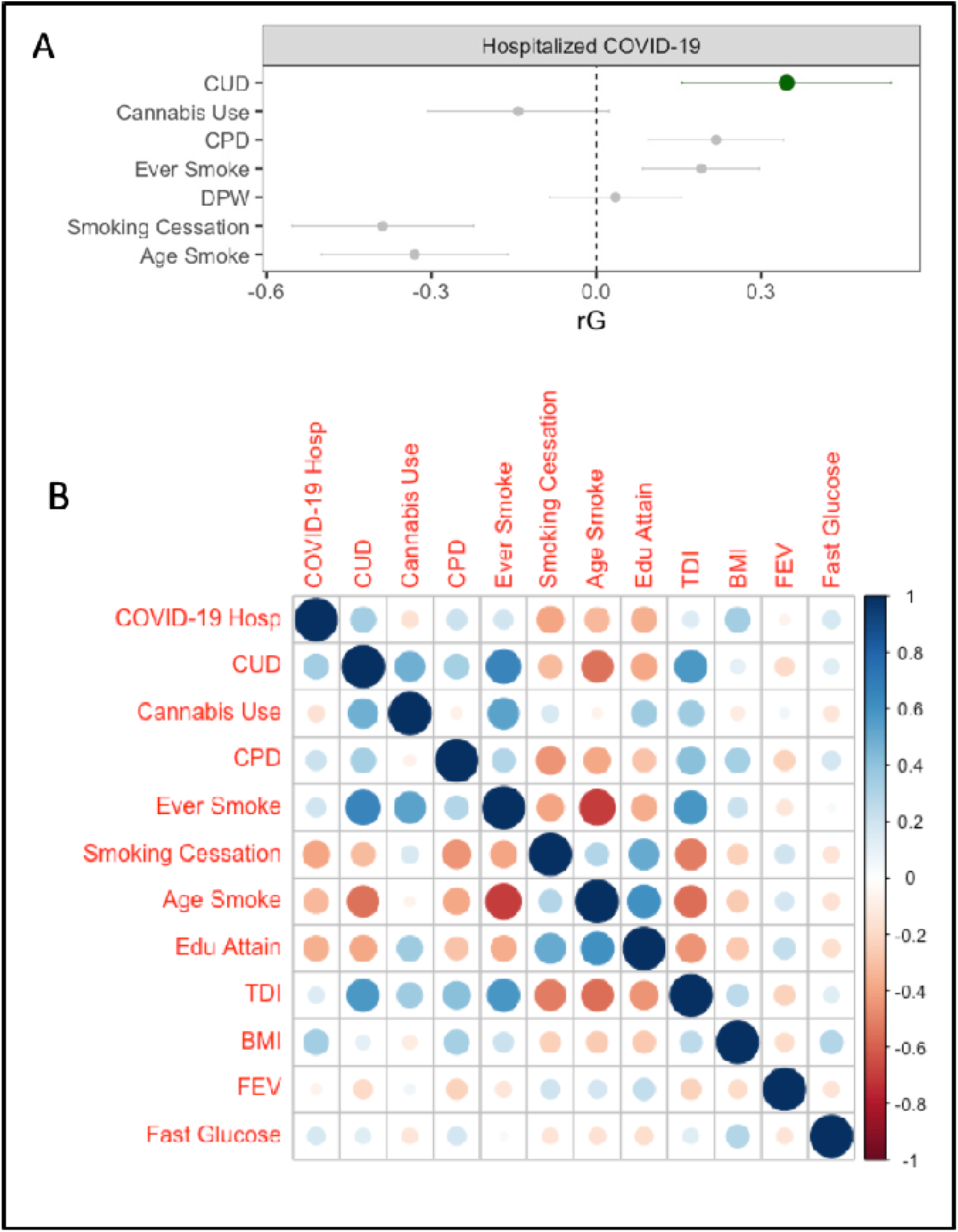
Genetic associations between Substance Use Phenotypes and COVID-19 Hospitalization. **A)** Genetic correlations between COVID-19 hospitalization and inhalation substance use phenotypes. Cannabis Use Disorder (CUD) is highlighted in green. CPD = Cigarettes per day, Age Smoke = Age of first becoming a regular smoker, Ever Smoke = Ever being a regular smoker, Cannabis Use = Any lifetime cannabis use. **(B)** Heat map of genetic correlations between all phenotypes included in the study. COVID-19 Hosp = COVID-19 hospitalization (Variable COVIDB2 from the COVID Host Genomics Consortium(4)). TDI = Townsend Deprivation Index, COVID-19 Hosp. = COVID-19 hospitalization, Edu Attain = Education attainment, BMI = Body Mass Index. FEV1 = Forced Expiration Volume for 1 second, Fast Gluc = Fasting Glucose.

**Table 1.** COVID-19 Genetic Correlations When Accounting for Potential Confounding Variables

| Smoking phenotypes | $\beta$ CUD | $\beta$ Smoking phenotype<sup>#</sup> | $\beta$ EA | $\beta$ TDI | $\beta$ BMI | $\beta$ FEV | $\beta$ Fast Glu |
| --- | --- | --- | --- | --- | --- | --- | --- |
| Cannabis Use | **0.566\*** | -0.362 | -0.004 | -0.109 | **0.288\*** | 0.093 | -0.001 |
| CPD | **0.358\*** | 0.044 | **-0.253\*** | **-0.256\*** | **0.292\*** | 0.075 | 0.032 |
| Age Smoke | **0.339\*** | -0.090 | **-0.220\*** | -0.265 | **0.296\*** | 0.068 | 0.034 |
| Smoking Cessation | **0.379\*** | **-0.345\*** | -0.140 | **-0.375\*** | **0.288\*** | 0.083 | 0.026 |
| Ever Smoke | **0.407\*** | -0.091 | **-0.260\*** | -0.221 | **0.311\*** | 0.075 | 0.026 |

**Note.** Standardized beta estimates for Cannabis Use Disorder (CUD) and smoking phenotypes were taken from a multiple regression parameterized in gSEM. <sup>#</sup>Smoking phenotypes were entered and tested separately to avoid multicollinearity amongst tobacco phenotypes. Each row represents genetic correlations with COVID-19 hospitalization from one model. The multiple rows indicate the separate models run substituting each tobacco phenotype (listed in the first column). The model was COVID hospitalization = CUD + Smoking Phenotype + BMI + Townsend Deprivation index + Educational Attainment + Forced Expiration + Fasting Glucose + error. The original GWAS accounted for standard GWAS covariates (age, sex, genetic principal components, etc). **Bold\*** represents $p < .05$ . MJ Use = Marijuana Use, CPD = Cigarettes per day, Age Smoke = Age started smoking regularly, Ever Smoke = Ever smoked regularly.

## Discussion

Our findings suggest that genetic liability to CUD is correlated with, and might increase risk for, COVID-19 hospitalization. The genetic correlation between CUD and COVID-19 hospitalization was independent of potential confounding variables (e.g., other substance inhalation phenotypes, cardio-metabolic traits, respiration, socioeconomic status indicators). On the other hand, genetic liability to lifetime cannabis ever-use showed a non-significant protective effect for COVID-19 hospitalization; divergent directionality of cannabis use and CUD has been observed for other phenotypes (e.g., BMI) (3). While highly speculative, it is possible that heavy and/or problematic cannabis use associated with CUD may increase COVID-19 severity, while any lifetime use may not be a marker of genetic susceptibility. Other putative mechanisms warrant attention in future research (e.g., potential shared effects on ACE2, inflammation). The use of population controls (i.e., individuals who may or may not have had COVID-19) in the COVID-19 hospitalization GWAS may have impacted association estimate precision and the predominant composition of European ancestry in constituent GWASs may limit generalizability to other ancestral populations. Finally, LCV models were unstable when only SNPs with MAF>0.05 were included and as such await replication (simulations suggest this may be attributable to reduced SNP density as opposed to MAF threshold; estimates of the GCP remained the same at 0.05 and were significant, but with wider standard errors). As the world prepares for surges in COVID-19, identifying putative risk factors associated with severe presentations may mitigate its worldwide impact. In contrast to anecdotal evidence and media reports (14) that cannabis may attenuate COVID-19, these data urge caution in heavy cannabis use during the COVID-19 pandemic.

## Materials and Methods

The following genome-wide association study (GWAS) summary statistics were used in analyses: CUD (n case=14,080, n control=343,726) (3) COVID-19 hospitalization (n case=6,492, n population control=1,012,809) (4), BMI (N= 795,640) (9), fasting glucose (N=58,074) (10), ever trying marijuana (N=184,765) (8), Forced Expiration Volume (11) (FEV; N= 272,338), Townsend deprivation index (11) (TDI; N= 336,798), education attainment (12) (N=766,345), cigarettes per day (N=263,954), ever a regular smoker (n=632,802), smoking cessation (N=312,821), and age of smoking initiation (N= 262,990; all smoking phenotypes are from the GSCAN consortium) (7). All GWAS summary statistics were generated from samples of genomically-confirmed European ancestry, with the exception of COVID-19 hospitalization which had 4.5% of the sample coming from other ethnic populations, and the remaining being of European descent.

### LDSCORE

Linkage Disequilibrium Score Regression (LDSC) (5) was used to estimate genetic correlations between traits using single nucleotide polymorphisms (SNPs) with minor allele frequencies (MAF)> 0,01, INFO > 0.90. Palindrome and multi-allelic single nucleotide polymorphisms (SNPs) as well as duplicate rs numbers and insertion/deletion polymorphisms were excluded before estimating genetic correlations. Single Nucleotide Polymorphisms (SNPs) with less than 1000 individuals were removed via the sample sizes of each GWAS (this was partially done to remove SNPs specific to anyone minority populations). LD score Beta weights and LD were pre-generated from 1000 Genomes European GWAS data included in the LDSC software download. LDSC regresses Chi-square statistics from the summary stats of GWAS on LD scores of the trait of interest (5). LD scores for each SNP are calculated via the sum of the variance explained by LD of that SNP with other SNPs (15). Genetic correlations were estimated using overlapping SNPs from filtered summary statistic files provided from GWAS and accounted for population stratification.

### Modeling Genetic Covariances from LDSCORE

Using the pairwise genetic correlation matrix, Genomic Structural Equation Modeling (gSEM) allows for estimating multivariate models with the genetic correlation matrix as input data. In this case, we can parameterize a multiple regression, with CUD genetics predicting COVID-19 vulnerability above and beyond other variables of interest. Smoking phenotypes were tested in separate models to avoid multicollinearity among tobacco phenotypes.

### Latent Causal Variable Analysis

LD scores can also be used to parameterize a quasi-experimental model. Building upon instrumental variable analyses and LDSR as applied to genomic data (often referred to as Mendelian Randomization), LCV models a latent “causal” variable which mediates the genetic correlation between two traits. This is quantified by the genetic causality proportion (GCP), an estimate of the degree to which each trait is correlated with the latent genetic variable, i.e., the extent to which each trait is potentially genetically causal (ranging from 0 reflecting no genetic causality to |1| indicating full genetic causality).

## Data Availability

Data are made available through the respective GWAS consortiums.

## Acknowledgments

ASH receives support from DA007261-17. CLM receives support from MH016880. EAW receives support from HD007289-30. ECJ receives support from AA027435-02. AA receives support from MH109532 and K02DA032573 Dr. Bogdan (AG052564, AA027827, DA046224). We also would like to acknowledge the COVID Host Genomics Consortium for their open data policies. All data are available through the PGC, GSCAN and COVID-HG consortiums. Conflict of interest disclosures: No disclosures were reported.

